# Predicting anti-cancer activity in flavonoids - a graph theoretic approach

**DOI:** 10.1101/2021.08.20.21262354

**Authors:** Simon Mukwembi, Farai Nyabadza

## Abstract

In drug design, there are two major causes of drug failure in the clinic. First, the drug has to work, and second, the drug should be safe. Identifying compounds that work for certain ailments require enormous experimental time and, in general, is cost intensive. In this paper, we are concerned with melanoma, a special type of cancer that affects the skin. In particular, we seek to provide a mathematical model that can predict the ability of flavonoids, a vast and natural class of compounds that are found in plants, in reversing or alleviating melanoma. The basis for our model is the conception of a new graph parameter called, for lack of better terminology, graph activity, which captures melanoma cancer healing properties of the flavonoids. With a superior coefficient of determination, *R*^2^ = 1, the new model faithfully reproduces anti-cancer activities of some known data-sets. We demonstrate that the model can be used to rank the healing abilities of flavonoids which could be a powerful tool in the screening, and identification, of compounds for drug candidates.

## 1 Introduction

In humans, a natural pigment, melanin, does not only provide a major defense mechanism against the ultraviolet light of the sun [17], but also determines the colour of the skin, hair and eyes [16]. The production of melanin is catalyzed by the enzyme, tyrosinase. The overproduction of melanin leads to undesirable skin conditions, such as hyperpigmentation and melanoma skin cancer, that have an effect on the quality of life. The cancer, melanoma, has increasingly become a common complaint among patients consulting with dermatologists [4].

Several agents, such as hydroquinone, corticosteroids, kojic acid and arbutin [2, 4], that reduce the production of melanin by inhibiting the activity of the catalyst, tyrosinase, are well known. Unlike natural tyrosinase inhibitors, which are generally considered to be cheap and free of harmful side effects, the existing traditional anti-tyrosinase agents suffer a legion of limitations such as high levels of toxicity, low stability, poor skin-penetration, and insufficient activity (see, for instance [4]). The effectiveness of a therapeutic agent is generally measured by the *inhibition concentration at* 50%, *IC*_50_, i.e., the quantity of the inhibitory agent required to inhibit the biological process, such as tyrosinase activity, by 50% [13]. The quantity *IC*_50_, is synonymous to the half-saturation constant in ecology. The half-saturation constant is defined as the resource availability at which half of the maximum intake is reached and it determines the outcome of models and may contribute to explain behavioural traits, life-strategies and species occurrence [8].

A well researched class of natural agents, flavonoids, commonly found in plants, have been reported to have anti-tyrosinase activity [11, 13, 16, 17]. Experiments to determine anti-tyrosinase activity of possible candidates for drug agents come with their own burdens. Among the burdens are high costs, laboratory experiment time, human efforts and the problems associated with animal sacrifice. Of particular concern however, as Hughes et al [6] puts it, drugs fail in the clinic for two main reasons; the first is that they do not work and the second is that they are not safe. Consequently, the development of mathematical models that can predict the effectiveness of compounds in inhibiting certain biological processes become handy.

In this paper, we will exploit graph theory to develop a model that predicts the anti-tyrosinase activity, i.e., *IC*_50_ values, of flavonoids. Our model can be used, not only to rationalise existing data, but also to predict new or unknown anti-tyrosinase activity in flavonoids.

## 2 Graphs

A *graph G* = (*V, E*) is a mathematical object which consists of a finite set *V* of elements called *vertices*, together with a set *E*, of 2-element subsets of *V*, called the *edges* of *G*. As early as 1875, Cayley (see for instance [1]), in his quest to enumerate chemical molecules called alkanes, he made an observation that molecules can be modelled by graphs where atoms are represented by vertices and two vertices are joined by an edge if the corresponding atoms are linked by bonds. This graph model became widely known as the *molecular graph*. The *degree*, deg *v*, of a vertex *v* of *G* is the number of edges incident with it. We say that *v* is an *end vertex* if its degree is 1. We will sometimes refer to the set of all end vertices as *external vertices* and call the set of all vertices of degree greater than 1 *internal vertices*. The *irregularity index, t*(*v*), of a vertex (introduced in [10] and applied to studies in chemistry in [5]) is defined as the number of neighbours of *v* with distinct degrees. The *distance d*_*G*_(*u, v*) between vertices *u* and *v* in *G* is defined as the length of a shortest path joining *u* and *v* in *G*. The *eccentricity* ec(*v*) of a vertex *v* of *G* is the distance between *v* and a vertex furthest away from *v* in *G*.

### 2.1 The graph parameters

Consider a connected graph *G* = (*V, E*) of order *n*, i.e., with *n* vertices. Over a hundred of graph parameters that are used to find relationships between the structure of a molecule and its physical properties in order to predict the physicochemical, biomedical, environmental and toxicological properties of a compound directly from its molecular structure are legion in the literature [3, 7, 12, 14, 15, 18]. It turns out that, to date, no relationships on the known parameters and anti-tyrosinase activity have been reported on. We create here a parameter that is broken down into two individual indices; one index specifically for external vertices, while the other one looks at internal vertices. We will then show that this new parameter can predict anti-tyrosinase activity.

Let *S* be the external vertices of *G*, i.e., the set of all end vertices in *G*. Let *Q* be the set of internal vertices of *G*. For a vertex *v* of *G*, the *score, s*(*v*), of *v* is the quantity

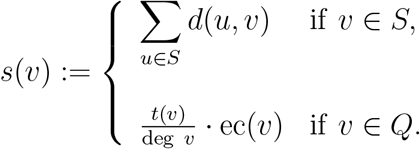

We define two invariants, *D*(*G*) and *ζ*(*G*), of *G* as follows:

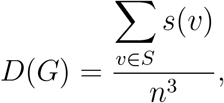

and

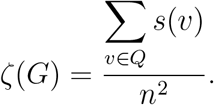

For lack of better terminology, we will call *D*(*G*) the *external activity* of *G*, and *ζ*(*G*), the *internal activity* of *G*, respectively. For instance, for the molecular graph *T* of the known standard tyrosinase inhibitor depicted in Figure 1, Taxfolin, we have

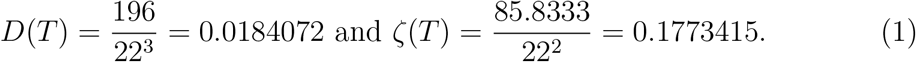

**Figure 1:**
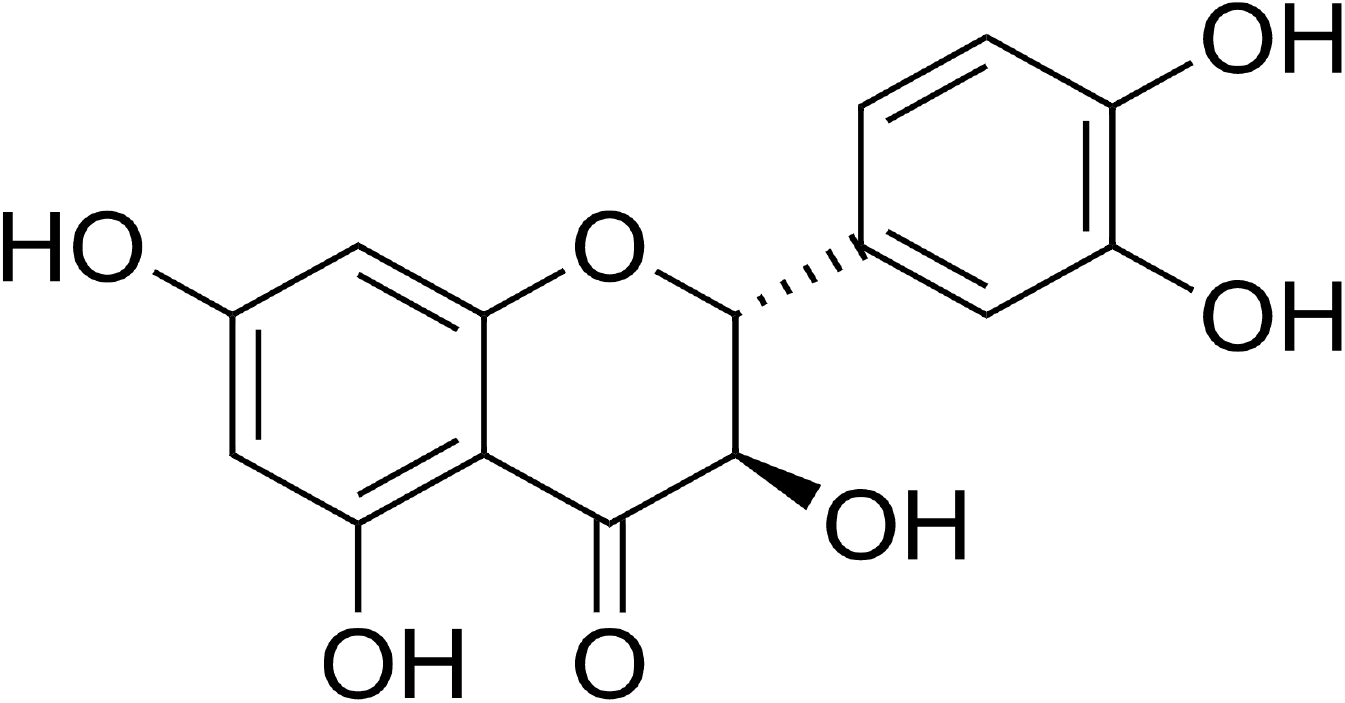
Tyrosinase inhibitor, Taxifolin (5,7,3’,4’-flavan-on-ol), also known as dihydroquercetin.

## 3 The model

Consider a flavonoid *F* whose molecular graph is *G*. We combine the internal and external activities of *G* to predict the anti-tyrosinase activity, i.e., *IC*_50_ value, of *F*. Table 1 presents flavonoids which were isolated from a plant in [17]. We will capitalise on this data-set to determine the values of the constants for our model, (2). We compute *D*(*G*) and *ζ*(*G*) for each of the flavonoid in Table 1. The graphs are shown in Figure 2.

**Table 1:** Data for nine flavonoids.

| **Flavonoid** | $D(G)$ | $\zeta(G)$ | $IC_{50}$ ( $\mu\text{M}$) |
| --- | --- | --- | --- |
| Dihydroquercetin-4'-methylether | 0.016848853 | 0.187145558 | 115 |
| Dihydroquercetin-7,4'-dimethylether | 0.015552662 | 0.1953125 | 162 |
| 5,7,3',5'-Tetrahydroxyflavanone | 0.014685239 | 0.155706727 | 423 |
| Blumeatin | 0.013429752 | 0.164600551 | 624 |
| Quercetin | 0.018407213 | 0.177341598 | 96 |
| Rhamnetin | 0.016931043 | 0.187775677 | 107 |
| Tamarixetin | 0.016931043 | 0.193761815 | 144 |
| Luteolin | 0.015117158 | 0.197656841 | 258 |
| Luteolin-7-methyl ether | 0.013899324 | 0.202479339 | 350 |

**Figure 2:**
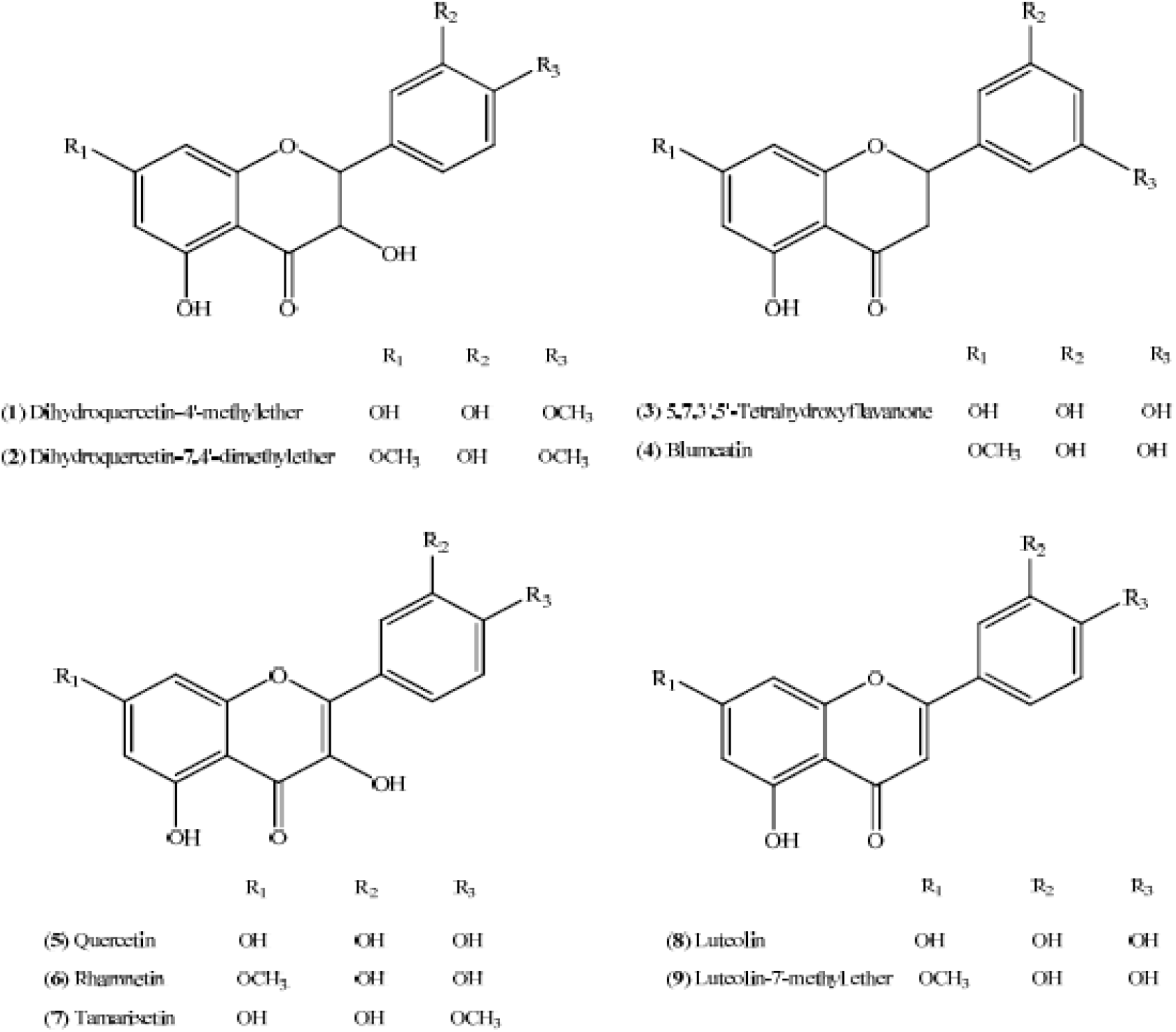
Molecular graphs of isolated flavonoids, Source: [17]

In Table 1, we present *D*(*G*) and *ζ*(*G*) of the flavonoids, together with their anti-tyrosine activities, which were determined in [17].

Given that the activity is driven by two invariants, *D*(*G*) and *ζ*(*G*), we propose a multivariable function to model the effects of the two invariants on the *IC*_50_ values. We propose a model of the form

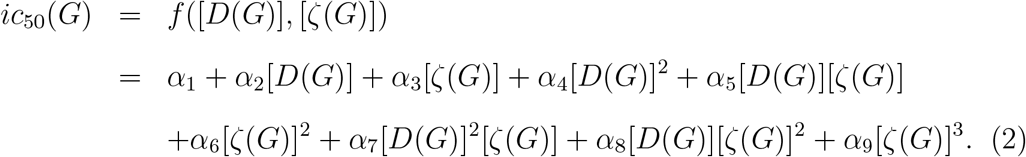

Fitting (2) to data given in Table 1, we obtain the values of the constants as

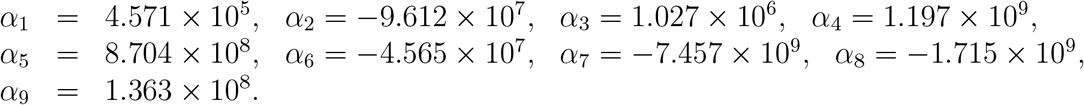

The given parameter values *α*_*i*_, *i* = 1, …, 9, produce a perfect fit of the model to the data. It is important to note that we resorted to the multivariable polynomial since it gives the best goodness of fit value measured by *R*^2^ or R-square. The strength of the relationship between a model and the dependent variables is measured by *R*^2^ ∈ [0, 1] or on a scale of 0 − 100%. We also note that the Curve Fitting Toolbox that we used in fitting our data to the model, gives a number of goodness of fit statistics for parametric models and in particular, the sum of squares due to error (SSE) and R-square. The summed square of residuals (SSE) is a measure of the total deviation of the response values from the fit to the experimental data. Our fit gives: SSE = 2.609 × 10^*−*19^ and *R*^2^ = 1.

A consideration of the contour map of Figure 3 gives the results in Figure 4. All the data points lie in the same contour colour indicating a perfect fit of the polynomial to the data.

**Figure 3:**
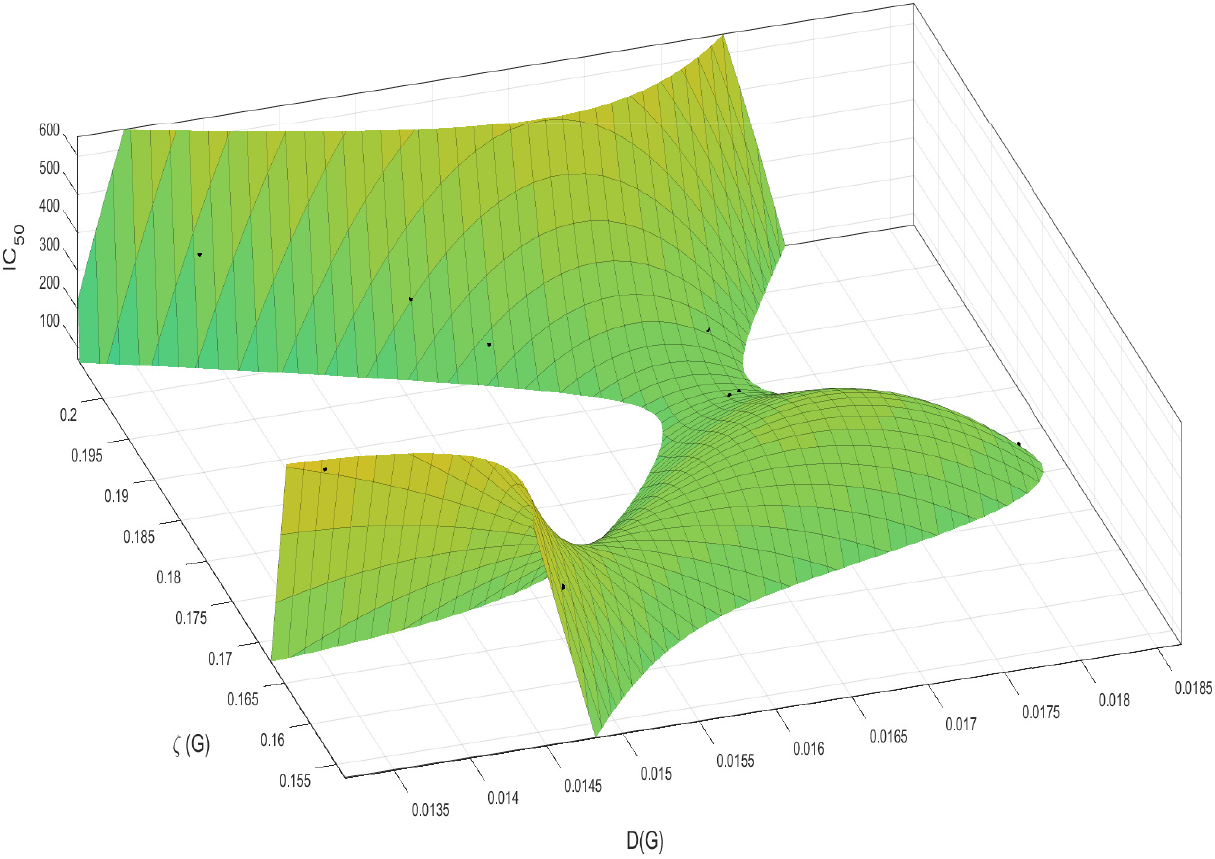
show the multivariable polynomial fit to the data.

**Figure 4:**
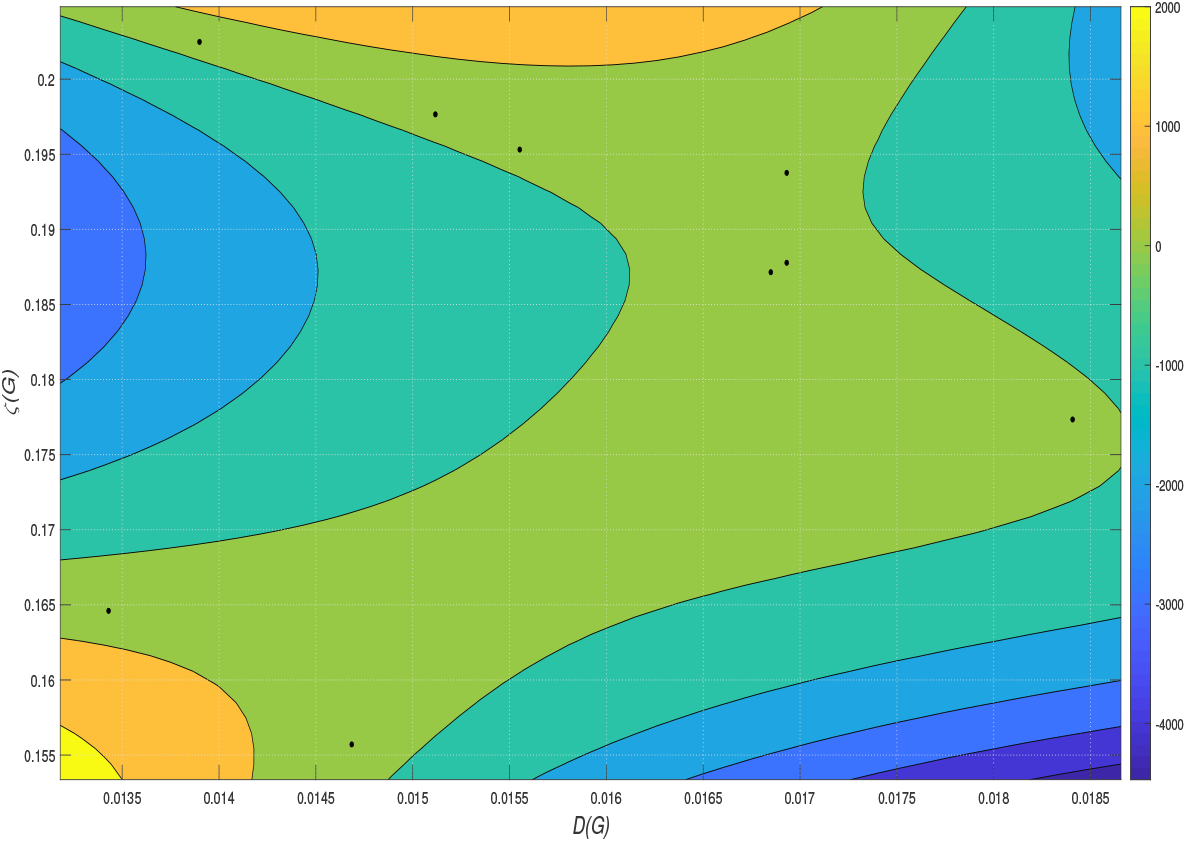
shows the contour plot of *D*(*G*) and *ζ*(*G*).

A plot of the residuals is shown in Figure 5. The residuals are found by looking at the difference between the data and the fit. The smaller the difference, the better the fit. The graph shows that the data points lie (almost) on the *D*(*G*)*ζ*(*G*)−trace, indicating very low values of the residuals.

**Figure 5:**
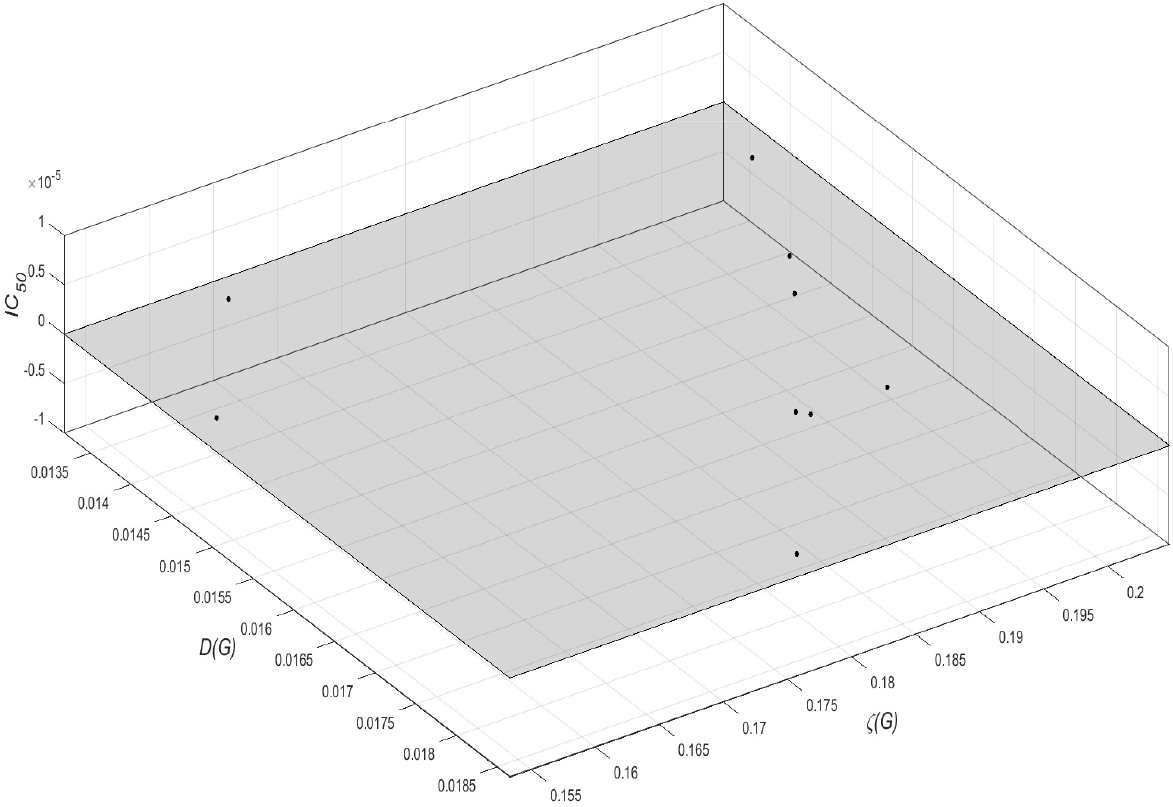
show the plot of the residuals, where residuals=data-fit.

## 4 Discussion and Applications

Our model has been developed, not only to rationalise existing data, but also to predict new or unknown anti-tyrosinase activity in flavonoids. However, for the sake of brevity and with such a very high goodness of fit, we use the newly developed model to rank the order of inhibition for 26 flavonoids. Additional flavonoids can, nevertheless, be added to the ranking list by using the model. The results are presented in Table 2.

**Table 2:**
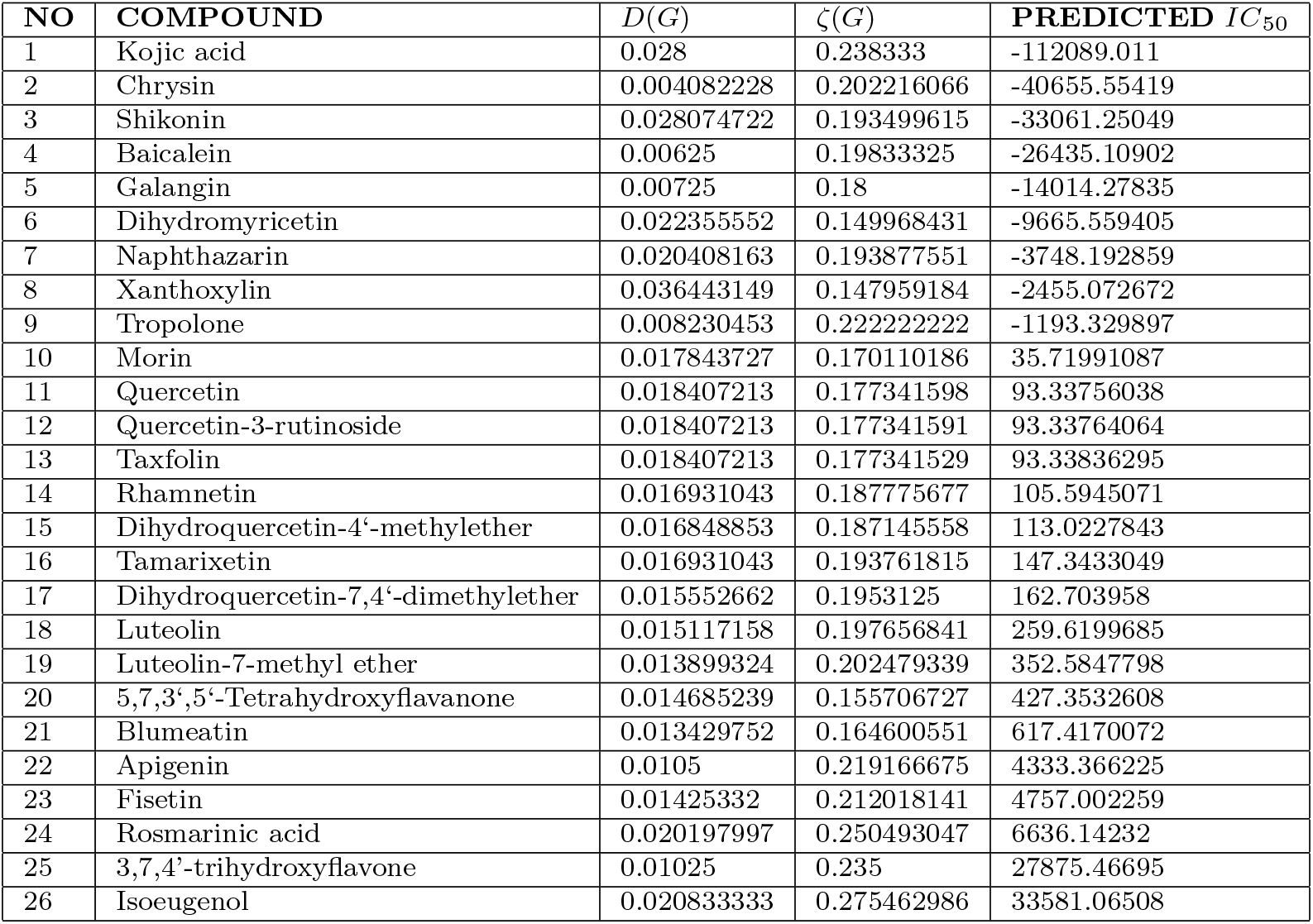
26 Flavonoids - rank order of inhibition.

Quite exciting is the fact that whilst the second compound, among the ranked flavonoids, chrysin, is commonly being used for bodybuilding, treating anxiety, inflammation, gout, erectile dysfunction, to mention but a few, it is appearing on the list as an excellent anti-tyrosinase agent which is only 3-fold weaker, and 34-fold stronger than the standard tyrosinase inhibitors, kojic acid and tropolone [2], respectively. Whilst these results on chrysin confirm Wu et al [19]’s findings that chrysin has potential use in skin photoprotection, Xie et al [20] found, in experimental work, that ‘chrysin had no effects on the (tyrosinase) enzyme’.

It is interesting to note that, through expanding the ranking list, our model can be used as a tool for identifying flavonoids that can be candidates for anti skin-cancer drug agents. This is critical in drug development as new drug agents get identified. Any identified agents can be tested using the model to determine their likelihood of use in fighting skin-cancer in the absence of laboratory tests.

Finally, we note here that it will be a worthwhile exercise to investigate the mathematical properties of the external, and internal activities of a graph, especially lower and upper bounds, and characterising the extremal graphs. Whilst external vertices play a significant role in influencing the anti-tyrosinase activity, the most dominant factor is the contribution made by the internal vertices. Each internal vertex *v*’s contribution is based on the distance between *v* and a vertex furthest away from *v*. This distance is then amplified by the irregularity of the neighbours of *v*.

## Data Availability

No data was used. What was used was obtained from literature and cited.

## References

[1] K.J. Burch, D.K. Wakefield and E.G. Whitehead, Jr, Boiling Point Models of Alkanes, MATCDY 47 (2003) 25–52.

[2] T.S. Chang, An Updated Review of Tyrosinase Inhibitors, International Journal of Molecular Sciences 10 (2009) 2440–2475.

[3] P. Dankelmann and S. Mukwembi, The Distance Concept and Distance in Graphs (I. Gutman, B. Furtula (Eds.)), Distance in Molecular Graphs Theory, Univ. Kragujevac, Kragujevac (2012) 3–48.

[4] M.M. de Freitas, P.R. Fontes, P.M. Souza, C.W. Fagg, E.N.S. Guerra, Y.K. de Medeiros Nobrega, D. Silveira, Y. Fonseca-Bazzo, L.A. Simeoni, M. Homem deMello, P.O. Magalhaes, Extracts of Morus nigra L. Leaves Standardized in Chlorogenic Acid, Rutin and Isoquercitrin: Tyrosinase Inhibition and Cytotoxicity, PLOS ONE (2016) 1–24.

[5] I. Gutman, Irregularity of molecular graphs, Kragujevac J. Sci. 38 (2016) 71—81.

[6] J.P. Hughes, S. Rees, S.B. Kalindjian and K.L. Philpott, Principles of early drug discovery, British Journal of Pharmacology 162 (2011) 1239-–1249.

[7] V. Sharma, R. Goswami, A. K. Madan, Eccentric connectivity index: A novel highly discriminating topological descriptor for structure-property and structure-activity studies. Journal of Chemical Information and Modeling 37 (1997) 273–282.

[8] C. Mulder, A. Jan Hendriks, Half-saturation constants in functional responses, Global Ecology and Conservation 2 (2014) 161–169

[9] M. Miyawaza and N. Tamura, Inhibitory compound of Tyrosinase activity from the sprout of Polygonum hydropiper L. (Benitade), Biological and Pharmaceutical Bulletin 30(3) (2007) 595-—597.

[10] S. Mukwembi, A note on diameter and the degree sequence of a graph, Applied Mathematics Letters 25 (2012) 175–178.

[11] R.J. Obaid, E.U. Mughal, N. Naeem, A. Sadiq, R.I. Alsantali, R.S. Jassas, Z. Moussa and S.A. Ahmed, Natural and synthetic flavonoid derivatives as new potential Tyrosinase inhibitors: a systematic review, RSC Advances 11 (2021) 22159—22198.

[12] M. Randic, A.T. Balaban, S.C. Basak, On structural interpretation of several distance related topological indices. Journal of chemical Information and Computer Sciences 41 (2001), 593–601.

[13] Y.B. Ryu, T.J. Ha, M.J. Curtis-Long, H.W. Ryu, S.W. Gal, and K.H. Park, Inhibitory effects on mushroom tyrosinase by flavones from the stem barks of Morus lhou (S.) Koidz, Journal of Enzyme Inhibition and Medicinal Chemistry 23(6) (2008) 922—930.

[14] S. Sardana, A.K. Madan, Application of graph theory: Relationship of antimycobacterial activity of quinolone derivatives with eccentric connectivity index and Zagreb group parameters. Match - Communications in Mathematical and in Computer Chemistry 45 (2002) 35–53.

[15] H.P. Schultz, Topological organic chemistry. 1. Graph theory and topological indices of alkanes. Journal of chemical Information and Computer Sciences 29 (1989) 227–228.

[16] B.D. Thach, V.Q. Dao, T.T.L. Giang, D.T. Cang, L.N.T. Linh, T.T. Ben, N.P.A. Uyen and N.K. Suong, Antioxidant and antityrosinase activities of flavonoid from blumea balsamifera (L.) DC. leaves extract, European Journal of Research in Medical Sciences 5(1) (2017) 1–6.

[17] N. Saewan, S. Koysomboon and K. Chantrapromma, Anti-tyrosinase and anticancer activities of flavonoids from Blumea balsamifera DC, Journal of Medicinal Plants Research 5(6) (2011) 1018–1025.

[18] H. Wiener, Structural determination of paraffin boiling points, Journal of the American Chemical Society 69 (1947) 17–20.

[19] N.L. Wu, J.Y. Fang, M. Chen, C.J. Wu, C.C. Huang, C.F. Hung, Chrysin protects epidermal keratinocytes from UVA- and UVB-induced damage. J Agric Food Chem 59(15) (2011) 391–400.

[20] L.P. Xie, Q.X. Chen, H. Huang, H.Z. Wang and R.Q. Zhang, Inhibitory effects of some flavonoids on the activity of mushroom tyrosinase. Biochemistry (Moscow) 68(4) (2003) 487–491.

